# Source terms for benchmarking models of SARS-CoV-2 transmission via aerosols and droplets

**DOI:** 10.1101/2022.02.11.22270844

**Authors:** Marc E.J. Stettler, Robert T. Nishida, Pedro M. de Oliveira, Léo C.C. Mesquita, Tyler J. Johnson, Edwin R. Galea, Angus Grandison, John Ewer, David Carruthers, David Sykes, Prashant Kumar, Eldad Avital, Asiri I.B. Obeysekara, Denis Doorly, Yannis Hardalupas, David C. Green, Simon Coldrick, Simon Parker, Adam M. Boies

**Affiliations:** Department of Civil and Environmental Engineering, Imperial College London, London, SW7 2AZ, UK; Department of Mechanical Engineering, University of Alberta, Edmonton, Alberta, T6G 2G8, Canada; Department of Engineering, University of Cambridge, Cambridge, CB2 1PZ, UK; Fire Safety Engineering Group, University of Greenwich, London, SE10 9LS, UK; Cambridge Environmental Research Consultants Ltd, 3 Kings Parade, Cambridge, CB2 1SJ, UK; AEROS Consultancy, Glasgow, G3 8SE, UK; Global Centre for Clean Air Research (GCARE), Department of Civil and Environmental Engineering, Faculty of Engineering and Physical Sciences, University of Surrey, Guildford GU2 7XH, UK; School of Engineering and Materials Science, Queen Mary University of London, London, E1 4NS, UK; Applied Modelling and Computation Group, Department of Earth Science and Engineering, Imperial College London, London, SW7 2AZ, UK; Department of Aeronautics, Imperial College London, London, SW7 2AZ, UK; Department of Mechanical Engineering, Imperial College London, London, SW7 2AZ, UK; Environmental Research Group, School of Public Health, Imperial College London, London, W12 0BZ, UK; Health and Safety Executive, Harpur Hill, Buxton, Derbyshire, SK17 9JN; Defence Science and Technology Laboratory, Porton Down, Salisbury, SP4 0JQ, UK

## Abstract

There is ongoing and rapid advancement in approaches to modelling the fate of exhaled particles in different environments relevant to disease transmission. It is important that models are verified by comparison with each other using a common set of input parameters to ensure that model differences can be interpreted in terms of model physics rather than unspecified differences in model input parameters. In this paper, we define parameters necessary for such benchmarking of models of airborne particles exhaled by humans and transported in the environment during breathing and speaking.

## 1 Introduction

Humans exhale particles made up primarily of respiratory fluid when breathing out, speaking, coughing, sneezing, singing and laughing and these particles may contain infectious pathogens (Stelzer-Braid et al., 2009; Yan et al., 2018). The size of exhaled particles spans several orders of magnitude and particle diameters range between 0.01 *−* 1000 µm (Bake, Larsson, Ljungkvist, Ljungström, & Olin, 2019). Historically, these particles have been classified in two categories by the infectious disease community: particles smaller than 5 µm in diameter are referred to as droplet nuclei or aerosols, whereas particles larger than 5 µm in diameter are classified as respiratory droplets (Milton, 2020; WHO, 2014). This somewhat arbitrary size classification implicitly refers to the transmission modes/mechanisms, namely airborne or droplet transmission, respectively. However, the connection between particle diameter (droplets vs aerosols) and the description of transmission mode/mechanisms (droplet vs airborne transmission) can lead to misunderstanding. For example, it is untrue in general that particles with diameter *>* 5 µm fall quickly onto a surface close to their source since these particles, particularly those *∼*5-10 µm in diameter, can be advected with ventilation flows over longer distances and remain airborne for longer periods. Consequently, Prather et al. (2020) recommend that aerosols and droplets are distinguished by a threshold of particle diameter of 100 µm, which more effectively separates their aerodynamic behavior, ability to be inhaled, and efficacy of interventions.

Particles are exhaled in a continuum of sizes and they rapidly change size depending on their environment, e.g. due to evaporation (Lieber, Melekidis, Koch, & Bauer, 2021). It is critical to understand the mechanisms of transport and deposition as a function of the size distribution of exhaled particles considering a range of external factors such as ventilation and air flows (Burridge et al., 2021). To that end, detailed experiments and models which accurately represent the relevant physics must be developed.

There is rapid advancement in approaches to modelling the fate of exhaled particles in different environments. These models have varying resolution and complexity in their representation of fluid flow and dispersion, and aerosol and droplet dynamics including evaporation, settling and transport(Chaudhuri, Basu, & Saha, 2020; de Oliveira, Mesquita, Gkantonas, Giusti, & Mastorakos, 2021; Shao et al., 2020; Wang, Wu, & Wan, 2020).

As these modelling approaches evolve, it is essential to understand their robustness in representing the different physical processes. An important aspect of this is an objective inter-model comparison so that any differences in results can be attributed to alternative implementation of the physics or purposeful differences in modelled conditions. With this paper, we provide a consolidated set of parameters for exhalation of particles that can be used by a range of modelling approaches as the basis for model inter-comparison.

Droplets and aerosols produced by violent exhalation events, such as coughing and sneezing, have been investigated and reviewed by several studies (Bourouiba, 2020; Bourouiba, Dehandschoewercker, & Bush, 2014; Mittal, Ni, & Seo, 2020; Yang, Lee, Chen, Wu, & Yu, 2007). Significant numbers of particles are also produced by breathing and speaking, activities which occur with greater frequency (Morawska et al., 2009). Under some circumstances, particularly in the case of presymptomatic or asymptomatic carriers who may not have symptoms of cough or sneezing, the cumulative amount of exhaled respiratory fluid as droplets and aerosols produced by high-frequency events such as breathing and speaking, may be greater than that due to low-frequency intermittent events (Dhand & Li, 2020). Furthermore, there remains uncertainty as to the importance of cough symptoms to transmission, with a recent study finding no association and that viral load, rather than symptoms, might be the predominant driver of transmission (Marks et al., 2021). We therefore focus on defining parameters for breathing and speaking.

Details omitted from the main text are included in the Supporting Information (SI) where referenced.

## 2 Model parameters and conditions

The set of parameters which characterize exhalation of particles and environmental conditions relevant to particle transport are shown in Table 1.

**Table 1:**
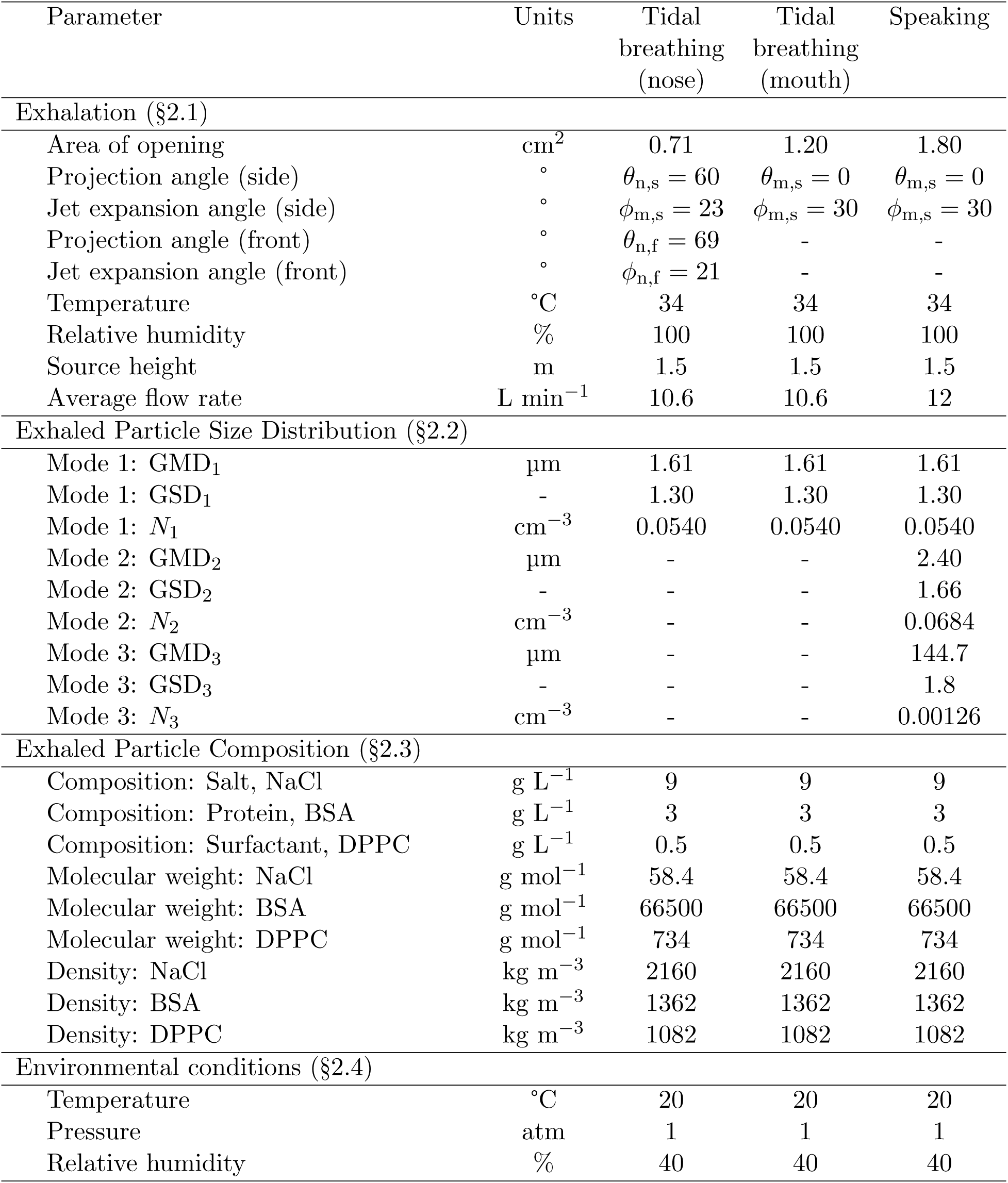
Parameters for modelling exhalation of particles.

### 2.1 Exhalation

Gupta, Lin, and Chen (2010) experimentally characterized various parameters associated with breathing and speaking; they measured gas flow rates, flow directions, and mouth and nose opening areas for 12 female and 13 male subjects. All subjects were healthy at the time of measurement and we note that there is a lack of literature on the potential effects of various symptoms of respiratory diseases on those parameters. The study documents significant variability amongst subjects and that flow rate is correlated to body surface area, which differs for males and females. The values listed in Table 1 represent nominal values for three cases of tidal (restful) breathing through the nose or mouth, and speaking.

Different models may have different requirements or constraints with regards to their representation of breathing. Breathing could be modelled as an unsteady phenomenon, or it may be more simplistically modelled as a constant flow rate. We have determined a self-consistent set of parameters for both approaches by conserving the total volume of exhaled air (and therefore the total number of exhaled particles). However, we note that this leads to different flow velocities at the mouth or nose opening as exhalation only occurs for approximately half of the breathing period.

The breathing air flow rate (*Q*; [L s^*−*1^]) can be modelled by a sinusoidal function (Gupta et al., 2010),

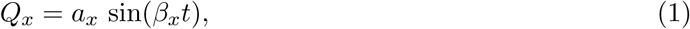

where *t* is time [s], the subscript *x* indicates either inhalation (in) or exhalation (out), *β*_*x*_ = *π* RF_*x*_*/*30 is a function of the respiratory frequency (RF; [min^*−*1^]), and *a*_*x*_ = *β*_*x*_ TV*/*2. The RF for inhalation (RF_in_) and exhalation (RF_out_) are given as functions of body height (*H*; [cm]) and body mass (*W* ; [kg]) by equations 7-10 in Gupta et al. (2010) and shown in the SI §SI-1. The tidal volume (TV; [L]) is given as

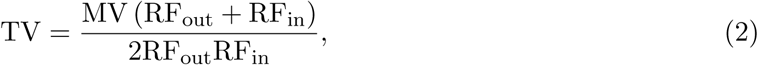

where the minute volume (MV; [L min^*−*1^]) is the volume of air exhaled in one minute (sometimes also referred to as the minute ventilation). The derivation of Eq. 2 is shown in the SI §SI-1. MV is correlated with the body surface area (BSA; [m^2^]) by MV = *c* × BSA. The constant *c* ([L min^*−*1^m^*−*2^]) is 5.225 and 4.634 for males and females, respectively (Gupta et al., 2010). The BSA can be estimated according to Gehan and George (1970),

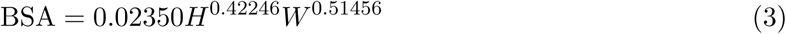

where *H* is height in cm and *W* is body mass in kg. Considering the average British male and female, who are 175.3 and 161.6 cm tall and weigh 83.6 and 70.2 kg, respectively (Office for National Statistics, 2010), we obtain *a*_in_ = 0.5956, *β*_in_ = 2.0629, *a*_out_ = 0.5215 and *β*_out_ = 1.8061 for males and *a*_in_ = 0.0.4794, *β*_in_ = 1.6722, *a*_out_ = 0.3991 and *β*_out_ = 1.3922 for females. Thus, the breathing flow rate (*Q*_breathing_; [L s^*−*1^]) over the cycle of inhalation and exhalation can be represented by a piecewise sinusoidal function with a period of *π/β*_in_ + *π/β*_out_,

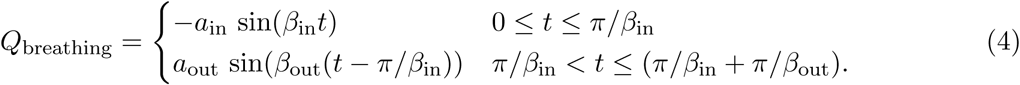

A graphical representation of *Q*_breathing_ for the average British male and female are shown in Figure 1. For the purposes of model comparison, we include *Q*_breathing_ for the average British male in Table 1.

**Figure 1:**
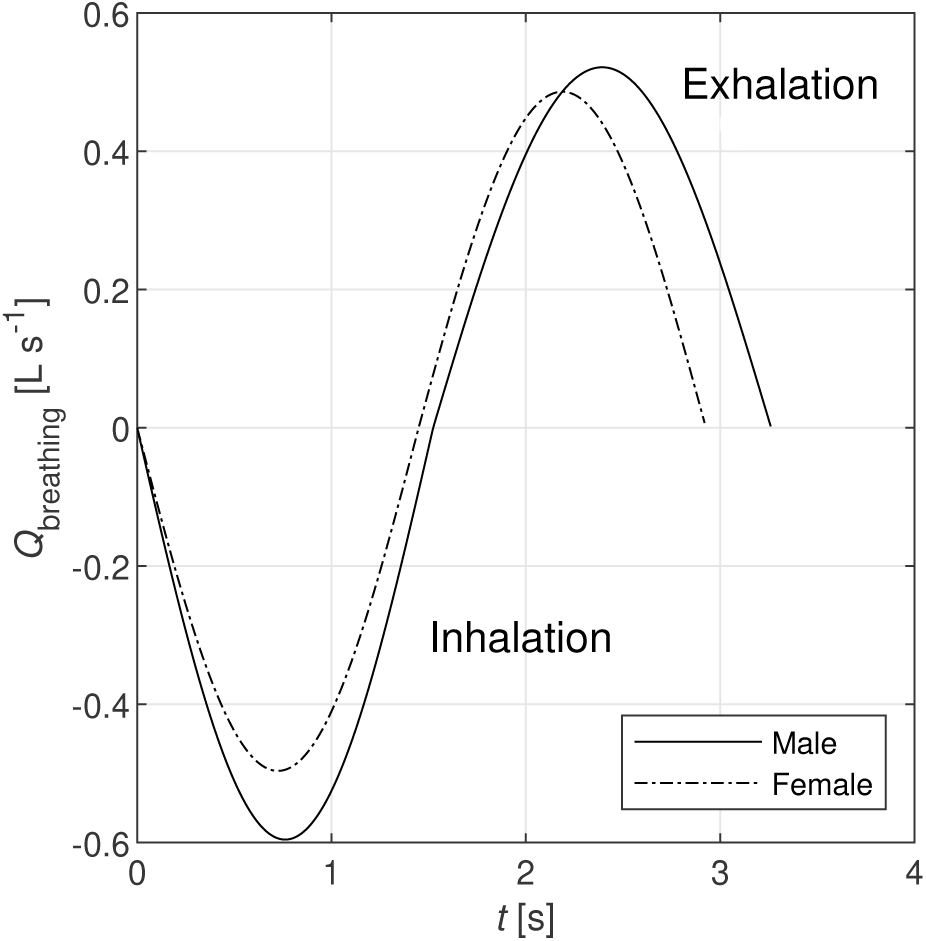
Graphical representation of the breathing flow rate.

Alternatively, the exhaled air flow rate may be modelled as a steady flow, in which case the average flow rate is obtained by dividing the total exhaled volume by the breathing period. Using the same values of *a*_out_ and *β*_out_, we obtain an average exhalation flow rate of 10.6 L min^*−*1^ (0.177 L s^*−*1^) and 8.3 L min^*−*1^ (0.139 L s^*−*1^) for the average male and female, respectively. These values are close to those recommended for representing breathing rates in risk assessments (Binkowitz & Wartenberg, 2001).

For speaking, the breathing pattern is not sinusoidal and varies significantly with the vocalization. A nominal average exhalation flow rate is 12 L min^*−*1^ (0.2 L s^*−*1^) for vocalizing passages of text (Gupta et al., 2010). While this is adequate for model comparison, we encourage readers to study the original reference for values that may be more representative of specific cases and to other literature that has measured the spread of exhalation flow rates for different individuals and vocalisations, which suggest that exhalation flow rates during singing are similar to those during speaking (Jiang, Hanna, Willey, & Rieves, 2016).

Nominal projection and spreading angles of the jets of exhaled air from the nose and mouth are also taken from Gupta et al. (2010) and they are shown graphically in Figure 2. For nose breathing, we suggest that it is appropriate to assume that the exhaled air flow is split equally between two nostrils. However we note that there is normally asymmetry in these flows due to anatomical, physiological and disease factors that shift and alternate the asymmetry over time (Hanif, Jawad, & Eccles, 2000).

**Figure 2:**
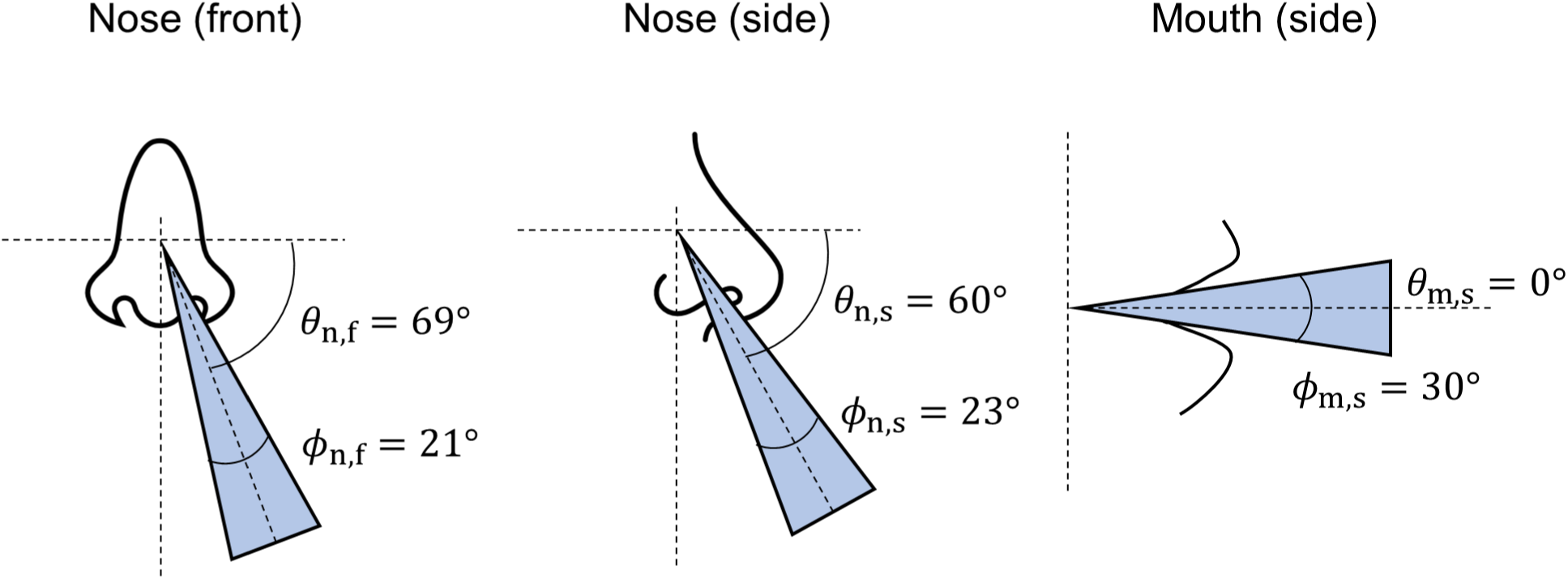
Graphical representation of jet projection (*θ*) and spreading (*ϕ*) angles.

### 2.2 Exhaled particle size distribution

The sizes of exhaled particles span several orders of magnitude and particle diameters range between 0.01 *−* 1000 µm (Bake et al., 2019). The earliest measurements of exhaled particle sizes relied on the microscopic analysis of droplet marks on slides placed in front of the mouth (Duguid, 1946) and these techniques are still used to estimate exhaled particle counts for particle diameters *>* 10µm (Johnson et al., 2011; Xie, Li, Sun, & Liu, 2009). Optical techniques have also been used to measure exhaled particles with diameters *>* 1 µm (Alsved et al., 2020; Chao et al., 2009). In studies using the droplet deposition and microscopy methods, it is common for the total number of particles counted within different size ranges to be reported, rather than the concentration of particles in exhaled breath and corrections are typically applied to the measured particle size distribution to account for artefacts such as evaporation or spreading of the droplets on the surface of the slide. To measure particles of diameter *<* 10 µm, a number of studies have relied upon measurements using the aerodynamic particle sizer (APS, Model 3321, TSI Inc.), which has a manufacturer-specified particle aerodynamic diameter detection range of 0.5 to 20 µm (Alsved et al., 2020; Asadi et al., 2019; Gregson et al., 2021; Johnson et al., 2011). These measurements are affected by the evaporation of water from the exhaled particles as they are expelled from the high humidity conditions in the body to the lower humidity of the experimental environment. The authors of these studies acknowledge that this process of droplet drying happens in the timescale of *∼* 1 s (Lieber et al., 2021) and that the measured size distribution is representative of the equilibrium size distribution. Johnson et al. (2011) applied a correction to account for the shrinkage of particles due to evaporation, whereas other studies have chosen not to correct for this process. Another important distinction between studies measuring particles in this size range is studies have either sampled a small fraction of the exhaled air flow (Asadi et al., 2019; Gregson et al., 2021) or have sampled the plume of exhaled air and corrected the measured concentration for plume dilution, as measured using a trace gas (e.g. water) (Johnson et al., 2011). A comparison of particle size distributions from different studies is shown in Figure 3 and details of the source of data for this plot can be found in the SI §SI-2.

**Figure 3:**
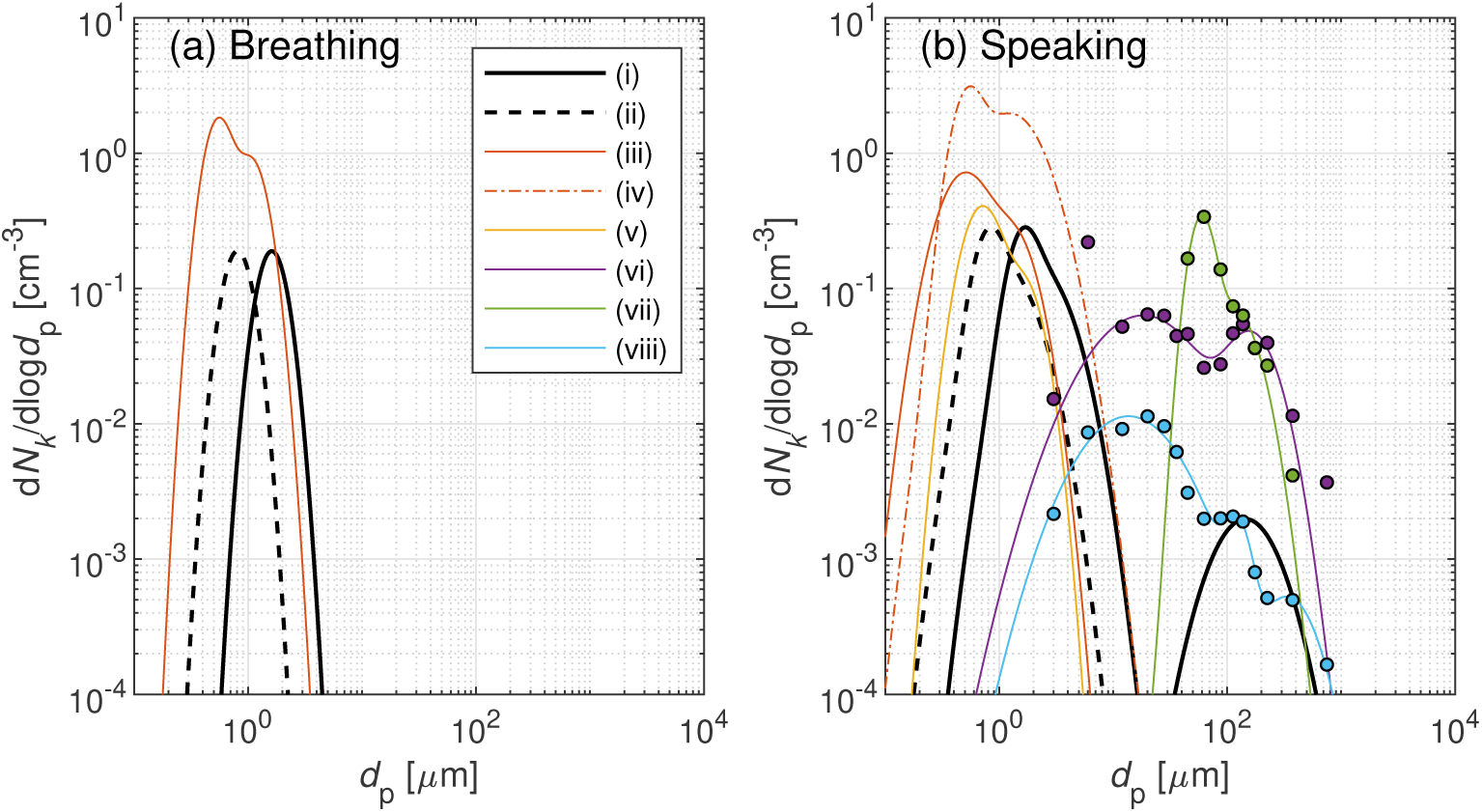
Exhaled particle size distributions resulting from (a) breathing and (b) speaking from (i) Johnson et al. (2011) corrected for particle shrinkage and representing the PSD at the mouth (*BLO model*), (ii) Johnson et al. (2011) not corrected for particle shrinkage (iii) Gregson et al. (2021) (70-80 dBA in (b)), (iv) Gregson et al. (2021) (90-100 dBA in (b)), (v) Asadi et al. (2019) (Fig. S10), (vi) Chao et al. (2009), (vii) Xie et al. (2009) and (viii) Duguid (1946). Parameters of lognormal distributions and further information on the sources of data are included in the SI §SI-2.

Johnson et al. (2011) reported that particles generated from breathing, speaking and coughing were present in a range of sizes, represented by distinct modes of a frequency distribution of particle diameters that spans from 0.1 to 1000 µm. They propose the *BLO model* for the size distribution of particles measured: bronchiolar (*B*), laryngeal (*L*) and oral (*O*) to represent the different locations in the airways believed to be the source of the aerosols.

A recent publication showed that patients admitted to hospital with COVID-19 exhaled similar aerosol size distributions to healthy patients when breathing, speaking and coughing (Hamilton et al., 2021).

#### Bronchiolar and laryngeal particles

Particle diameters from the first two modes, bronchiolar and laryngeal, were found to range from at least 0.5 to 5 µm, both with median diameters of order 1 µm using on-line measurement techniques using the APS and after correction for evaporation by assuming a shrinkage factor of 0.5 (Johnson et al., 2011). The evaporation-corrected size distribution represents the initial particle size distribution at the mouth and can be compared to the uncorrected equilibrium size distribution in Figure 3. Recently, Asadi et al. (2019) and Gregson et al. (2021) reported equilibrium particle size distributions for breathing and speaking. For speaking, both studies report significant variability with respect to the loudness of vocalisation and amongst individuals. As shown in Figure 3, these two studies are in good agreement with the uncorrected size distribution from Johnson et al. (2011) with respect to median diameters. However, the three studies span approximately an order of magnitude in concentration and the size distributions from Asadi et al. (2019) and Gregson et al. (2021) appear to have a larger spread (i.e. geometric standard deviation). The difference in concentration between studies is likely within the range of variation due to vocalization, loudness and individual variability, however it is possible that sampling and data processing differences may also contribute.

While we focus here on breathing and speaking, we acknowledge that there are recent studies reporting particle size distributions for singing (Alsved et al., 2020; Gregson et al., 2021). While singing is found to increase the number concentration of exhaled particles relative to speaking, the increase is small relative to the changes associated with increased loudness (Gregson et al., 2021).

#### Oral particles

Johnson et al. (2011) reported that the oral mode of particles measured during speaking were larger than 10 µm in diameter and all contained food-dyed saliva, demonstrating that those particles originated from the mouth. This observation of the presence of food-dye is in agreement with Duguid (1946) and Xie et al. (2009), and data from these two studies are also shown in Figure 3. We have also included the optical measurements from Chao et al. (2009) in Figure 3 and it is evident that there is significant variation in the magnitude, mode and spread of size distributions for oral particles. These differences may be attributed to differences in measurement techniques, vocalizations and variability amongst individuals. It is beyond the scope of this paper to review these differences in detail, however we note the need for further studies that compare different measurement approaches, for example by conducting simultaneous measurements using different techniques of the same exhaled aerosol, and the interested reader is referred to the following additional references (Almstrand et al., 2010; Loudon & Roberts, 1967; Papineni & Rosenthal, 1997; Stadnytskyi, Bax, Bax, & Anfinrud, 2020). We recommend that the oral particle size distribution for speaking is treated as more uncertain than the bronchiolar and laryngeal modes. The parameters for the size distributions from different studies are included in the SI §SI-2 to enable model sensitivity studies. There is limited evidence of exhaled aerosols with diameters *>* 10µm as a result of singing.

#### Parameter specification

The discussion above indicates that there is significant variability in exhaled particle concentration and size distribution due to respiratory activity and individual variability. For the purposes of model comparison, we adopt the *BLO model* (Johnson et al., 2011), corrected to represent the particle size distribution at the mouth (series (i) in Figure 3), as the basis of the terms included in Table 1. We note that this particle size distribution is representative of the mean for the group of healthy volunteers in that study and is therefore not predictive of a single person as inter- and intra-person variability is of the order of measured concentration itself or greater (Johnson et al., 2011).

For breathing, only the *B* mode is included. For speaking, the size distribution of exhaled particles is the sum of the three *B, L* and *O* lognormal distribution modes (Johnson et al., 2011),

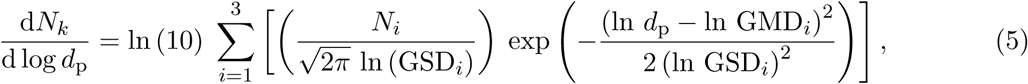

where *d*_p_ is the particle diameter [µm], *N*_*i*_ is the total number concentration [cm^*−*3^] of each mode *i*, GMD_*i*_ is the geometric mean diameter [µm] of each mode *i*, and GSD_*i*_ is the geometric standard deviation of each mode *i*. Each mode may be characterized by only three parameters: GSD_*i*_, GMD_*i*_, and *N*_*i*_ as given in Table 1. The particle size distribution for breathing and speaking is shown in Figure 4(a).

**Figure 4:**
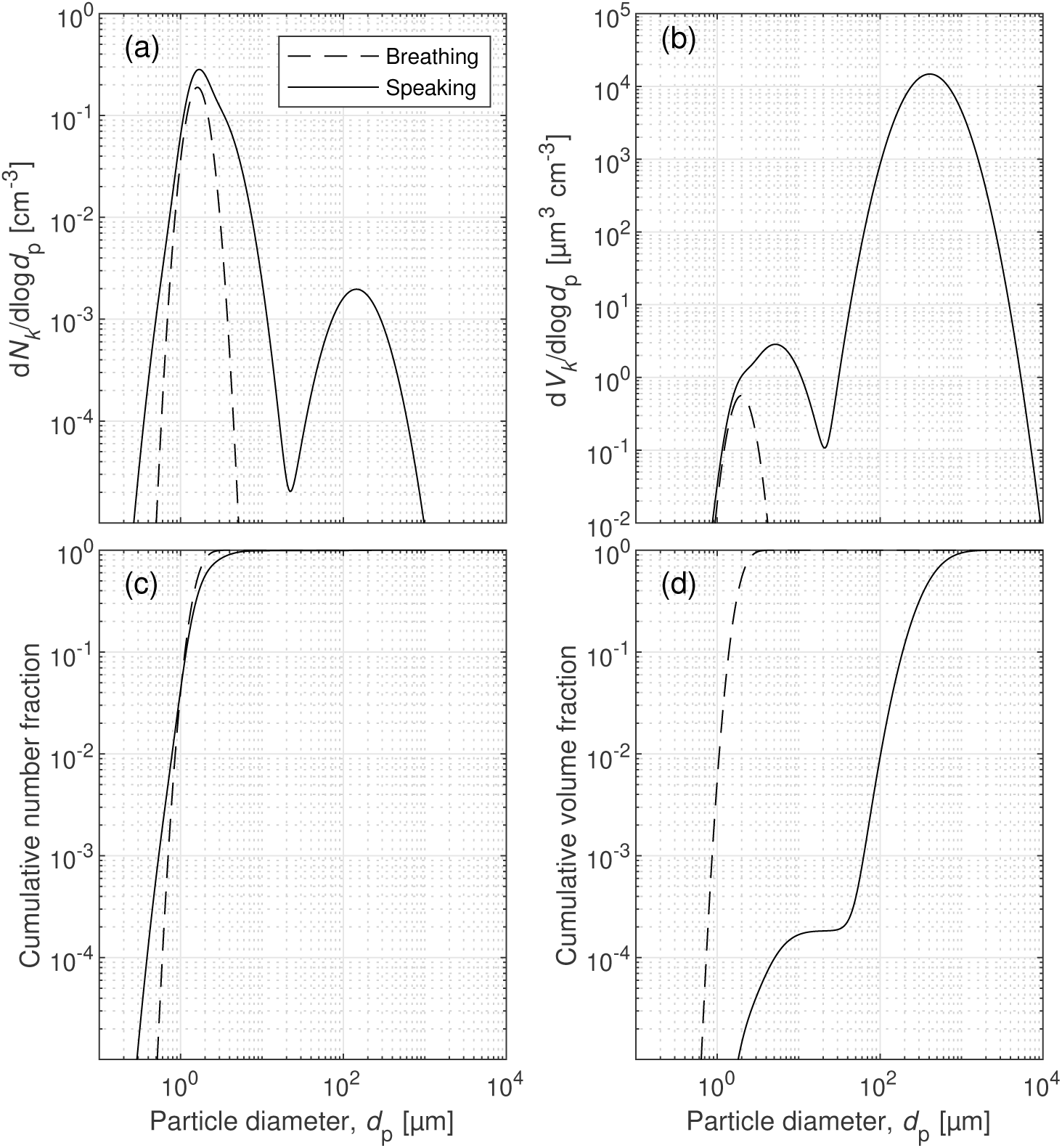
(a) Number and (b) volume weighted particle size distributions, and cumulative fractions of (c) particle number and (d) volume as a function of particle diameter for breathing and speaking.

The notation d*N*_*k*_/d log *d*_p_ represents the number concentration in each bin of particle diameters (d*N*_*k*_) normalized by a bin width that is constant in log space, i.e. d log *d*_p_ = log(*d*_p,*k*+1_*/d*_p,*k*_), where *k* represents a discretization of the *d*_p_ space. Note that log here refers to the base 10 logarithm and Eq. 5 is preserved from Johnson et al. (2011). Therefore, the absolute number concentration of particles of a given bin of particle diameters (d*N*_*k*_; [cm^*−*3^]) is calculated as,

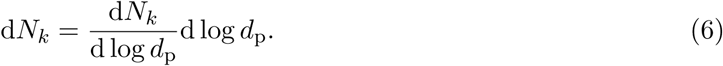

In the context of exhalation, it is important to consider both number and volume of exhaled particles. The volume of particles of a given diameter represented as a concentration [µm^3^cm^*−*3^], assuming all particles are spherical, is given by

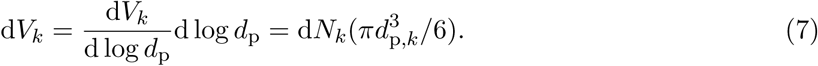

The volume weighted particle size distribution and cumulative number and volume fractions are shown in Figure 4(b-d) for breathing and speaking. The total number concentration, *N*, of particles is 0.054 cm^*−*3^ for breathing and 0.1237 cm^*−*3^ for speaking and the total volume concentration, *V*, is 0.1608 µm^3^ cm^*−*3^ for breathing and 9.4637 10^3^ µm^3^ cm^*−*3^ speaking, summarized in Table 2.

**Table 2:** Estimates of concentrations and emission rates of particles.

| Parameter | Units | Breathing | Speaking |
| --- | --- | --- | --- |
| Nominal average flow rate: $\bar{Q}$ | $\text{cm}^3 \text{s}^{-1}$ | 176 | 200 |
| Exhaled number concentration: $N$ | $\text{cm}^{-3}$ | 0.054 | 0.124 |
| Exhaled volume concentration: $V$ | $\mu\text{m}^3 \text{cm}^{-3}$<br>($\text{mL cm}^{-3}$) | 0.161<br>($1.61 \times 10^{-13}$) | $9.46 \times 10^3$<br>($9.46 \times 10^{-9}$) |
| Avg. number emission rate: $\bar{E}_N$ | $\text{s}^{-1}$ | 9.50 | 24.7 |
| Avg. volume emission rate: $\bar{E}_V$ | $\mu\text{m}^3 \text{s}^{-1}$<br>($\text{mL s}^{-1}$) | 28.3<br>($2.83 \times 10^{-11}$) | $1.89 \times 10^6$<br>($1.89 \times 10^{-6}$) |

The release rate of particle number (*E*_*N,k*_; [s^*−*1^]) or volume (*E*_*V,k*_; [µm^3^s^*−*1^]) for a given particle diameter is calculated as the product of the particle number or volume concentration and the exhaled flow rate, i.e.

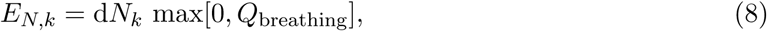

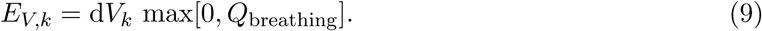

For example, considering a nominal average flow rate of 12 L min^*−*1^ (200 cm^3^ s^*−*1^) for vocalizing passages of text (Gupta et al., 2010) and exhaled number, *N*, and volume, *V*, concentrations for speaking yields emission rates of *Ē*_*N*_ = 24.7 s^*−*1^ or volume *Ē*_*V*_ = 1.89 10^6^ µm^3^ s^*−*1^ (1.89×10^*−*6^ mL s^*−*1^). Estimates of particle emission rates during breathing and speaking are summarized in Table 2, highlighting that speaking produces an estimated 6.7 10^4^ times larger volume of fluid than breathing alone, primarily from the oral mode of droplets (typically larger than 10-50 µm) originating from the mouth.

### 2.3 Exhaled particle composition

Exhaled particles are multi-component droplets comprising water, salts, proteins and surfactants (Effros et al., 2002; Vejerano & Marr, 2018; Veldhuizen & Haagsman, 2000). Once exhaled from the nose or mouth, these particles are exposed to a rapidly changing relative humidity (RH) within the exhaled breath from approximately 100% to ambient conditions. The combination of the droplet composition and ambient temperature and relative humidity will influence the evaporation rate and therefore affect settling times of a single respiratory droplet (Lieber et al., 2021; Marr, Tang, Van Mullekom, & Lakdawala, 2019; Walker et al., 2021). As a multi-component droplet with non-volatile solutes evaporates, the evaporation rate may change throughout the process due to an increase in concentration of solutes in the liquid, as well as other physico-chemical transformations (Vejerano & Marr, 2018). The resulting size of the droplet, represented by a characteristic diameter, after it has come into equilibrium with the ambient conditions not only determines its settling time (de Oliveira et al., 2021; Marr et al., 2019; Walker et al., 2021; Xie et al., 2009), but also its fate in the respiratory system should it be inhaled by an individual (ICRP, 1994; Madas et al., 2020; Milton, 2020). When considering the whole range of sizes found in respiratory releases (Fig. 4), the combined effect of relative humidity and composition may result in up to an order-of-magnitude difference in the total amount of suspended mass of a droplet cloud of different compositions (de Oliveira et al., 2021).

For the purposes of model comparison, we suggest a four-component droplet composition consisting of 9 mg mL^*−*1^ of NaCl, 3 mg mL^*−*1^ of protein (bovine serum albumin, BSA), and 0.5 mg mL^*−*1^ of surfactant (dipalmitoylphosphatidylcholine, DPPC) in water. This protein concentration is representative of the composition of nasal surface airway fluid (Gould & Weiser, 2001) and this simplified composition is comparable to concentrations in simulated lung fluid (Hassoun, Royall, Parry, Harvey, & Forbes, 2018; Kumar et al., 2017; Walker et al., 2021), and has been used in a previous modelling study (de Oliveira et al., 2021). The concentration of each component, together with their respective molecular weight and density (Haynes, 2013; Nagle & Wilkinson, 1978; NCBI, 2020; Tanford, 1961) are given in Table 1. Properties of water for modelling the dynamics of particles including evaporation are readily available from e.g. Green and Perry (2019). The three other components (i.e. NaCl, protein, and surfactant) are not volatile at typical ambient conditions due to their significantly higher molecular weights and melting points (ChemSpider, 2020; Michnik, 2003). We note that when modelling the dispersal of virus within respiratory fluid (c.f. §3), the contribution of virus particles to the bulk composition of the particle is negligible for typical viral loads.

### 2.4 Environmental conditions

The temperature and relative humidity of ambient air significantly affects the fate of exhaled particles, in terms of the rate of evaporation of water from droplets (de Oliveira et al., 2021; Redrow, Mao, Celik, Posada, & Feng, 2011), and, while not not explicitly relevant to defining source terms, the inactivation rates of enveloped viruses (K. Lin & Marr, 2020; Marr et al., 2019; Morris et al., 2021).

Guidelines for different indoor environments are published by various regulatory bodies. For example, guidelines for ward spaces and intensive care units in hospitals, set design air temperature and relative humidity ranges between 20-24°C and 30-60%, respectively (Beggs, Kerr, Noakes, Hathway, & Sleigh, 2008) and similar guidelines exist for schools in different countries (Education and Skills Funding Agency (ESFA), 2018; Mohamed, Rodrigues, Omer, & Calautit, 2021; S, E, Y, & E, 2014).

Empirical studies on indoor temperature and relative humidity in different environments suggest that these can vary with the seasons and that there is variability between buildings used for the same purpose. In three hospitals in the US, temperatures were measured to within the recommended range of 20-24°C, however relative humidity was consistently below 40% in all locations (Quraishi, Berra, & Nozari, 2020). In two hospitals in France, average temperatures and humidity ranged from 19-27°C and 16-70%, respectively, across all seasons (Baurès et al., 2018). For low-income households in the UK in winter, median indoor temperature and relative humidity were found to be 19°C [14-23°C, 5th to 95th centile range] and 43% [32-60%] in living rooms, respectively (Oreszczyn, Ridley, Hong, Wilkinson, & Group, 2006), with significant variability by season and dwelling type (McGill, Oyedele, & McAllister, 2015). For dwellings in the US, median indoor temperature and relative humidity were found to be 20°C [18-27°C, range] and 48% [23-71%], with seasonal variations in relative humidity (Nguyen, Schwartz, & Dockery, 2014). In industrial settings, there may be indoor conditions that are specific to the activity and setting, e.g. meat processing (Günther et al., 2020), and standards and outdoor conditions will have an effect in different climatic regions (De Vecchi, Candido, de Dear, & Lamberts, 2017; Wu, Cao, & Zhu, 2018).

The empirical evidence suggest that temperature and relative humidity span the range of 20-24°C and 30-60%, respectively. For model comparison we propose an ambient temperature of 20°C and a relative humidity of 40%, which are included in Table 1. Given the importance of these parameters, we would encourage researchers to present results for the ranges of 15-30°C and 30-60%, as a minimum.

## 3 Pathogens in exhaled particles

There is limited evidence for the amounts of pathogens possibly contained in particles exhaled by different respiratory activities and significant variability among different types of pathogens, therefore we do not include values for concentrations of pathogens in our set of parameters for exhaled particles. However, considering the recent focus on modelling transmission of SARS-CoV-2, below we discuss the data for SARS-CoV-2 to help readers make more informed judgments on appropriate viral load values for their modelling efforts.

### 3.1 Prevalence of SARS-CoV-2 in indoor air

At the time of writing, the viral load and infectivity of SARS-CoV-2 in exhaled particles of different sizes has not been well established (Dabisch et al., 2020). Gene copies^1^ of SARS-CoV-2 ribonucleic acid (RNA) have been detected by polymerase chain reaction (PCR) analyses of samples of indoor air gathered in a range of (mostly clinical) settings (Zhang & Duchaine, 2020), including in aerosols smaller than 5 µm (Chia et al., 2020; Feng et al., 2021; Liu et al., 2020). In indoor air, the concentrations of SARS-CoV-2 RNA reported in particles smaller than 5 µm are of order 1*×*10^*−*5^ (Liu et al., 2020) to 1*×*10^*−*3^ (Chia et al., 2020; Feng et al., 2021; Ong et al., 2021) gene copies per cm^3^ of sampled air.

Importantly, modellers must note that the number of SARS-CoV-2 gene copies detected by PCR quantifies sub-sections of the viral RNA sequence and is therefore not equal to the number of infectious viruses present. However, based on a range of clinical samples (e.g. nasopharyngeal swabs), the likelihood of detecting infectious SARS-CoV-2 by viral culture methods is correlated with number of gene copies reported where RNA viral loads greater than 10^5^-10^6^ gene copies/mL (corresponding to Ct*<∼*24-25) and higher are typically required to demonstrate infectivity of a clinical sample containing SARS-CoV-2 (Bullard et al., 2020; Cevik et al., 2021; Huang et al., 2020; Kim et al., 2021; Y.-C. Lin et al., 2021; Singanayagam et al., 2020; van Kampen et al., 2021; Wölfel et al., 2020). To date, cycle thresholds for the air samples that detect SARS-CoV-2 RNA are very often *>*30 and even *>*35, implying air samples are often not likely to culture (Zhang & Duchaine, 2020). Of attempts to demonstrate the infectivity of SARS-CoV-2 suspended in field samples of indoor air by viral culture methods (Döhla et al., 2020; Ong et al., 2021; Santarpia et al., 2020; Yamagishi et al., 2020; J. Zhou et al., 2020), there has been limited evidence of viral replication or cytopathic effects (CPE) (Lednicky et al., 2020, 2021; Santarpia et al., 2020).

Plaque assay, a cell culture method, when performed on samples with higher viral loads than typically found in air samples (e.g. naso-pharyngeal swabs), enables quantification of the number of infectious viruses capable of forming plaques in a cell monolayer, called plaque forming units (PFU). These PFUs may be used in dose-response models to estimate infection risk in humans (as done for SARS, for example (Watanabe, Bartrand, Weir, Omura, & Haas, 2010)). Syrian hamsters inoculated by the intranasal route were infected with a dose of as low as 14 PFUs and the minimum infectious dose may be lower in humans (Y.-C. Lin et al., 2021). Since there is insufficient data on the possible load of infectious viruses in air samples, it is appropriate to estimate a possible range based on the number of gene copies detected. A ratio of RNA gene copies (N Gene) to PFUs of *∼*160,000:1 was found using almost 500 clinical samples (including naso-pharyngeal swabs, sputum, saliva, and fomites) from 75 patients. A ratio of *∼*10,000:1 was reported when using a more homogeneous virus that can be harvested from culture in a laboratory setting (Y.-C. Lin et al., 2021), in line with other studies (Plante et al., 2021). Therefore, roughly assuming an RNA:PFU ratio of 10,000:1, air concentrations of 1*×*10^*−*3^ (Chia et al., 2020; Feng et al., 2021; Ong et al., 2021) gene copies per cm^3^ of sampled air would correspond to 1*×*10^*−*7^ PFU per cm^3^ of air (or one PFU in ten cubic meters of air). Measurements of viral prevalence in indoor air includes many variables depending on the situation. To model viral exhalations, it is preferred to use empirical data from direct measurements of viruses contained in exhaled air, or data of viral load contained in the respiratory tract fluid that is exhaled.

### 3.2 Prevalence of SARS-CoV-2 in air directly exhaled by infected persons

SARS-CoV-2 RNA has been detected in exhaled breath condensate (EBC), where participants’ exhaled breath is cooled and its contents are condensed into solution for analysis, without resolving the exhaled particle size distribution (Ding et al., 2021; Feng et al., 2021; Ma et al., 2020; Ryan et al., 2021; L. Zhou et al., 2021). Some studies report that concentrations in excess of *∼*10^*−*1^ gene copies per cm^3^ of exhaled breath are possible, calculated based on their PCR results for EBC and the volume of air sampled (Ma et al., 2020; L. Zhou et al., 2021). Recent studies use a sampling apparatus which separates bioaerosols into ‘coarse’ (*>*5 µm) and ‘fine’ (*≤*5 µm) fractions to compare exhalations from breathing, speaking and coughing (Coleman et al., 2021) or assess the performance of masks (Adenaiye et al., 2021) for the amount of SARS-CoV-2 exhaled. These studies report significantly lower RNA exhalation rates than Ma et al. (2020) reported for EBC. More data of direct measurements of exhalations are needed to provide more confidence in models of virus exhalations, however these studies provide insights to quantify virus exhalaton rates (Adenaiye et al., 2021; Coleman et al., 2021).

Studies haven’t yet attempted to quantify indoor air samples relative to exhaled breath samples for the same participants, and comparisons between studies are subject to variabilities in viral load of patient, variant type, room air ventilation rates (and, designed vs actual ventilation rates), variance in expiration rates based on patient (e.g. patient coughing vs breathing). A value of 10^*−*3^ gene copies per cm^3^ for room air (Chia et al., 2020; Feng et al., 2021; Ong et al., 2021) and 10^*−*1^ gene copies per cm^3^ for exhaled breath (Ma et al., 2020; L. Zhou et al., 2021) would suggest a reasonable dilution ratio of 100, but this relation may be coincidental and more systematic sampling is required.

### 3.3 Prevalence of pathogens in respiratory fluid

Caution must be exercised if estimating viral load from samples of fluid extracted directly from the respiratory tract (e.g. naso-pharyngeal swabs). Aerosols are plausibly generated from small airways (Johnson & Morawska, 2009; Malashenko, Tsuda, & Haber, 2009), airway walls (Moriarty & Grotberg, 1999), larynx (Johnson & Morawska, 2009), and mucosalivary fluid from the mouth (Bourouiba, 2021a; Johnson et al., 2011) by a range of mechanisms. Measurements of viral load in respiratory fluid span several orders of magnitude, they change over the course of the disease and they can be different depending on the source of respiratory fluid (Singanayagam et al., 2020; Walsh et al., 2020; Wölfel et al., 2020).

To date, many studies have assumed a constant concentration of viruses in the fluid that composes the exhaled particles across the continuum of particle sizes (Stadnytskyi et al., 2020) to assess relative risk rather than absolute risk of disease transmission associated with the modelled scenarios. Given that assumption, considering the cumulative volume fractions in Figure 4 show the vast majority of respiratory fluid by volume is in the oral mode, it is expected the vast majority of viral RNA detected would be found in the oral mode. However, recent data from the studies discussed in Section 3.2 question this assumption. Coleman et al. (2021) reported from direct measurements of breathing, speaking and coughing, that 85% of the detected RNA copies were found in the fine (*≤*5 µm) aerosol fraction compared with the coarse (*>*5 µm) aerosol fraction. Comparable results, where similar or more viral RNA is found in the fine aerosol mode, have been found for influenza (Leung et al., 2020; Milton, Fabian, Cowling, Grantham, & McDevitt, 2013; Yan et al., 2018), and these results have substantial implications for the relative importance of short versus long range transmission. However, the viral RNA possibly carried by the largest droplets may not be detected if they, for example, drop into the walls of the cone of the Gesundheit-II apparatus used in Coleman et al. (2021) and are not retrieved. Cheng et al. (2021) discussed the discrepancy between measurements of viral exhalations with other measurements of aerosol/droplet volumes as a function of particle size citing a possible gradient in viral load throughout the respiratory tract.

In light of this recent evidence, in the SI §SI-3 we propose a method for scaling a viral load in the B and L modes relative to the O mode of the exhaled particles so that researchers can test their models in the limit where viral load in fine aerosols is significantly higher. We present this in general terms such numerical values can be updated as more evidence becomes available. Taking the measurements from Coleman et al. (2021), where 85% of the viral load to be in the particles with diameter less than 5 µm, we calculate that the viral load in the B and L modes would need to be 6*×*10^5^ times higher than the viral load in the O mode.

There is a critical need improve the empirical data for the viral load in different particle sizes. Evidence from swab samples reported by Tu et al. (2020) showed tongue swabs, perhaps representing the oral mode, contained generally lower viral RNA loads than NP swabs, perhaps representing the B and L models, though by 1-2 orders of magnitude, not 4-5.

#### Correcting conversions for volumes of respiratory fluid

We do not recommend directly using clinical data of gene copies per mL reported for swab samples that have been diluted into another fluid. For example, naso-pharyngeal swab samples are submerged and transported in viral transport media (typically in 3 mL of transport media) (Wölfel et al., 2020). Subsequent measurements of viral RNA by PCR could be reported in gene copies per mL of transport media or gene copies per swab. However, the exact volume of respiratory fluid sampled on a given swab is unknown. While this is roughly of order 0.1 mL, the volume collected depends on the type of swab, practitioner and properties of the fluid. The dilution correction is therefore not well known and furthermore elution of viruses from the swab may be incomplete (Warnke, Warning, & Podbielski, 2014). More discussion may be found in Roque et al. (2021) which points out that if the average NP swab collects and releases 50 µL of nasal secretions and is stored 3 mL of transport media, the original sample is diluted 60:1. Then, volumes extracted from the total solution for analyses by PCR must be correctly accounted. These conversions may be estimated for modelling purposes, however it must again be noted that the viral load may be different in different regions of the respiratory tract.

### 3.4 Experimental data needed for estimation of viral loads in aerosols and droplets

There are significant complexities of gathering experimental data relevant to disease transmission. Considering only aerosol sampling, it is difficult to gather size-resolved measurements of viral load in a controlled manner. For particle diameters larger than *∼*10 µm, competing transport phenomena (e.g. convection, gravity, inertial impaction) affect sampling, which may introduce bias in the reported results. Depending on the bioaerosol sampling method, the range of particle sizes sampled must be carefully considered. For example, the smallest particles *<*0.3 µm are inefficiently captured in a BioSampler (Lindsley et al., 2017). Furthermore, for there to be an infection risk, the pathogen must be viable at the time of exhalation and must survive in the exhaled aerosol particles or droplets. The survival of viruses and bacteria in aerosols and droplets is highly dependent on the environmental conditions, such as the relative humidity, temperature and exposure to light (Dabisch et al., 2020; K. Lin & Marr, 2020; Marr et al., 2019). Therefore, it is critical that both gene copies and attempts to culture the virus are reported in measurements along with resolution of viral load as a function of particle size.

Additionally, more measurements of exhaled particle size distributions are needed. Specifically, since the particle sizes emitted vary by several orders of magnitude (*∼* 0.01-1000 µm), more data are needed from instruments which complement one another to capture the entire size ranges of aerosols and droplets for the same exhalatory activities (Bourouiba, 2021b). These data which are available in a controlled setting are critical to reconcile with viral exhalation rates described above, which are arguably more difficult to gather. By combining data of viral exhalations and aerosol/droplet exhalations, more accurate assessments of relative risk of different modes of transmission in specific scenarios are possible.

## 4 Summary and recommendations

There is rapid advancement in approaches to modelling the fate of exhaled particles in different environments. As these modelling approaches evolve, it is important that each model implementation can be verified by comparison with others, and that any differences in results can be attributed to incomplete specification or alternative implementation of the physics. With this paper, we provide a consolidated set of parameters for exhalation of particles that are appropriate to be used by a range of modelling approaches as the basis for model inter-comparison and benchmarking.

In reporting results, details of all physical and mathematical models should be provided along with a description of the modelled scenario including a diagram, dimensions, and all boundary conditions. It is necessary to resolve particle transport (and deposition) as a function of particle diameter, therefore distributions of both number concentrations and volume concentrations (as shown in Figure 4) should be reported as a function of time and spatial location relative to the particle source (e.g. in vertical and horizontal cross sections). By reporting distributions of particles by volume, models for viral load within each particle may be readily applied to model virus dispersal and deposition, allowing relative assessments of risk relative rather than absolute assessments of risk.

While this paper focuses on defining a set of terms for model benchmarking, there is significant person-to-person variability in exhaled air flows, exhaled particle distributions and composition, and, perhaps most significantly, in viral load. The evidence base for the statistical distribution of these parameters within the population is incomplete; different studies typically have small sample sizes and are not often directly comparable, for example due to different vocal activities and measurement methods. Thus, there is insufficient evidence to quantify the modal, mean or median parameter values within the population. We therefore strongly encourage modelers to account for the sensitivity of their results to these uncertainties: exhaled air flow variability could be quantified using distributions of body height and weight (Gupta et al., 2010; Jiang et al., 2016); a number of different measured exhaled aerosol distributions are presented in the SI §SI-3; different representations of respiratory droplet composition could be used (Edwards et al., 2021; Vejerano & Marr, 2018; Walker et al., 2021); and, the large range in viral load discussed in Section 3 must be accounted for in any attempt to quantify the absolute risk of transmission.

There remain a significant number of outstanding questions related to airborne transmission of pathogens. Modelling the fate of exhaled particles, when implemented with careful verification of methods and experimental validation, can help to understand possible transmission pathways and inform efforts to mitigate transmission.

## Data Availability

All data produced in the present study are available upon reasonable request to the authors.

## Acknowledgements

This work was motivated and enabled by the work and discussions co-led by Professors Paul Linden and Christopher Pain under Task 7 (Environmental and aerosols transmission) of the Royal Society’s ‘Rapid Modelling of the Pandemic project’ (RAMP).

## Footnotes

1 Studies commonly report gene copies of a target gene (e.g. N gene of SARS-CoV-2) which are converted from cycle thresholds (Ct) by calibration for a given PCR system. Cycle thresholds represent the number of amplification cycles used to detect a target gene by PCR, where lower Ct values correspond to higher numbers of gene copies. Since Ct are platform dependent, it is preferred to compare among studies using gene copies determined by a standard calibration procedure.

