## Supplemental Information for "Source terms for benchmarking models of SARS-CoV-2 transmission via aerosols and droplets"

### Supporting Information

### SI-1 Tidal and minute volumes

The tidal volume (TV; [L]) is the volume of air exhaled in one breath. The minute volume (MV; [L min$^{-1}$]) is the volume of air exhaled in one minute. Therefore, MV can be written as the product

*

of the TV and the number of breaths in a minute ($n$; [$\text{min}^{-1}$]).

$$\text{TV} = \frac{\text{MV}}{n}. \tag{SI-1}$$

The sinusoidal model of breathing represented by Eq.1 in the main text defines the period ($T$; [s]) of the inhalation or exhalation as,

$$T_x = \frac{2\pi}{\beta_x}, \tag{SI-2}$$

where the subscript $x$ indicates either inhalation (in) or exhalation (out). The respiratory frequency [$\text{min}^{-1}$] is the number of inhalations or exhalations per minute,

$$\text{RF}_x = \frac{60}{T_x} = 30\frac{\beta_x}{\pi}. \tag{SI-3}$$

Equivalently,

$$\beta_x = \frac{\pi \text{RF}_x}{30}. \tag{SI-4}$$

To find the number of exhalations in a minute, $n$, consider that the total period of the cycle of inhalation followed by exhalation is the sum of half the period of inhalation and half the period of exhalation,

$$T = \frac{\pi}{\beta_{\text{in}}} + \frac{\pi}{\beta_{\text{out}}}. \tag{SI-5}$$

Substituting for $\beta_x$ gives,

$$T = 30\left(\frac{\pi}{\text{RF}_{\text{in}}} + \frac{\pi}{\text{RF}_{\text{in}}}\right). \tag{SI-6}$$

The number of exhalations in a minute is therefore the number of times the period can complete in 60 seconds,

$$n = \frac{60}{T} = \frac{2\text{RF}_{\text{out}}\text{RF}_{\text{in}}}{\text{RF}_{\text{out}} + \text{RF}_{\text{in}}}. \tag{SI-7}$$

Therefore, the tidal volume can be expressed as,

$$\text{TV} = \frac{\text{MV}}{n} = \frac{\text{MV}\left(\text{RF}_{\text{out}} + \text{RF}_{\text{in}}\right)}{2\text{RF}_{\text{out}}\text{RF}_{\text{in}}}. \tag{SI-8}$$

Correlations between the respiratory frequency and an individual's height ($H$; [cm]) and body mass ($W$; [kg]) are given by Gupta, Lin, and Chen (2010) for males and females separately,

$$\text{RF}_{\text{in,male}} = 55.55 - 0.3286H + 0.2602W, \tag{SI-9}$$

$$\text{RF}_{\text{out,male}} = 77.03 - 0.4542H + 0.2373W, \tag{SI-10}$$

$$\text{RF}_{\text{in,female}} = 46.43 - 0.1885H, \tag{SI-11}$$

$$\text{RF}_{\text{out,female}} = 54.47 - 0.2548H. \tag{SI-12}$$

### SI-2 Particle size distribution parameterisations

Table SI-1 includes the parameters of the exhaled particle size distributions from different studies included in Figure 3 of the main text. Parameters from Johnson et al. (2011) and Gregson et al. (2021) are taken directly from each of these articles. The data shown in Figure S10 of Asadi et al. (2019), representing the particle size distribution measured for one person, was obtained via personal communication and we fitted a bimodal lognormal distribution. We have assumed that the number concentration reported in all three studies can be directly compared and that differences in sampling procedures can be accounted for. In the case of Johnson et al. (2011), the concentration was corrected for plume dilution using water as a tracer gas, and for Gregson et al. (2021) and Asadi et al. (2019) we assumed that the reported concentration is undiluted as a result of these experiments only sampling a small proportion of the exhaled air flow rate into the APS.

Data from Chao et al. (2009) corresponds to the particle number concentration estimated using the laser volume (L.V.) method described in their study (Table 5 therein) and this was converted to $\mathrm{d}N_k/\mathrm{d}\log d_\mathrm{p}$ by diving the number counts by the width of each size bin. The data point corresponding to a particle diameter of 6 µm was treated as an outlier and not included in the fitting procedure. To estimate particle number concentration using the data from Xie, Li, Sun, and Liu (2009) (Table 5 therein) and Duguid (1946) (Table 3 therein), it was necessary to divide the reported particle counts by the exhaled volume of air and we used a similar approach to that described by Johnson et al. (2011): we assumed an average exhaled flow rate for talking of 12 $\mathrm{L}^{-1}$ (c.f. Table 1 of the main text) (Gupta et al., 2010) and that it would take one second to count each number from 1 to 100, i.e. 100 s in total, leading to 20 L of exhaled air. We then calculated $\mathrm{d}N_k/\mathrm{d}\log d_\mathrm{p}$ by dividing the number concentration in each size bin by the width of the size bin. For Duguid (1946), we fitted a tri-modal lognormal distribution as this gave a better fit to the data. However, this does not suggest that these three modes represent the *BLO* modes described by Johnson et al. (2011) and the data from Duguid (1946) and Xie et al. (2009) are representative of the oral mode due to the presence of food dye, which had been inserted in to the mouth prior to the experiment, in the observed particles.

Regarding the discrepancies in reported size distribution of oral particles, it is important to note that Duguid (1946), Xie et al. (2009) (deposition and microscopy) and Chao et al. (2009) (interferometric Mie imaging) used a single measurement technique that spanned the majority of the relevant particle size range for oral particles. While Xie et al. (2009) also included an aerosol spectrometer, they found that this instrument was not able to consistently measure exhaled droplets and therefore only measurements from the deposition method were reported. In contrast, Johnson et al. (2011) used a combination of APS and deposition measurements and they note that their droplet deposition technique was insensitive to particles with diameter less than 20 µm. This may explain the apparent spectral gap in the *BLO* model and the significantly lower concentration relative to the other studies near 20 µm, as shown in Figure 3 of the main text. We recommend that the oral size distributions for speaking are treated as highly uncertain and we encourage sensitivity studies. Further experimental work on full spectrum droplet measurement from respiratory activities should be a priority area for future research.

Table SI-1: Parameters of the particle size distributions shown in Figure 3 of the main text. Note that the modes labelled 1-3 do not necessarily correspond to the *BLO* modes described by Johnson et al. (2011).

| | | | Mode 1 | | | Mode 2 | | | Mode 3 | | |
|---|---|---|---|---|---|---|---|---|---|---|---|
| Ref | Legend | Activity | $N$ | GMD | GSD | $N$ | GMD | GSD | $N$ | GMD | GSD |
| | | | $cm^{-3}$ | µm | - | $cm^{-3}$ | µm | - | $cm^{-3}$ | µm | - |
| Johnson et al. (2011) | (ii) | Breathing | 0.0540 | 0.81 | 1.30 | 0.0000 | | | | | |
| | (ii) | Speaking | 0.0540 | 0.81 | 1.30 | 0.0684 | 1.20 | 1.66 | 0.00126 | 144.67 | 1.80 |
| | (i) | Breathing (at mouth) | 0.0540 | 1.61 | 1.30 | | | | | | |
| | (i) | Speaking (at mouth) | 0.0540 | 1.61 | 1.30 | 0.0684 | 2.40 | 1.66 | 0.00126 | 144.67 | 1.80 |
| Gregson et al. (2021) | (iii) | Speaking 70-80 dBA | 0.3540 | 0.50 | 1.58 | 0.1000 | 1.34 | 1.48 | | | |
| | (iv) | Speaking 90-100 dBA | 0.7490 | 0.53 | 1.32 | 1.2230 | 1.28 | 1.78 | | | |
| | - | Singing 70-80 dBA | 0.3950 | 0.52 | 1.32 | 0.4950 | 1.14 | 1.70 | | | |
| | - | Singing 90-100 dBA | 1.0000 | 0.55 | 1.26 | 2.0930 | 1.27 | 1.82 | | | |
| | (iii) | Breathing | 0.4940 | 0.55 | 1.29 | 0.2660 | 1.07 | 1.32 | | | |
| Asadi et al. (2019) | (v) | Speaking | 0.1540 | 0.71 | 1.42 | 0.0370 | 1.65 | 1.38 | | | |
| Chao et al. (2009) | (vi) | Speaking (1-100) | 0.0647 | 18.30 | 2.56 | 0.0219 | 163.06 | 1.58 | | | |
| Xie et al. (2009) | (vii) | Speaking (1-100) | 0.0774 | 59.37 | 1.25 | 0.0342 | 110.19 | 1.55 | | | |
| Duguid (1946) | (viii) | Speaking (1-100) | 0.0108 | 13.65 | 2.40 | 0.0004 | 120.91 | 1.27 | 0.00028 | 342.85 | 1.65 |

### SI-3 Viral load in the B & L modes relative to the O mode

The number of gene copies present in a volume of air ($C$; [copies cm$^{-3}$]) may be quantified by the particle volume concentration ($V$; [µm$^3$ cm$^{-3}$]), e.g. from Table 2 in the main text, multiplied by the viral load ($\lambda$; [copies mL$^{-1}$]) of the fluid particles contained in that volume of air, i.e.

$$C = V\lambda. \tag{SI-13}$$

For lack of evidence, previous studies (e.g. Stadnytskyi, Bax, Bax, and Anfinrud (2020)) have assumed a constant viral load for all exhaled particles. However, since the generation mechanisms of the B, L, and O modes are different and may be made up of fluid from different parts of the respiratory system, we can allow for different viral loads of gene copies within the fluid of each mode, and write that the total number of gene copies is the sum of the contribution from each mode,

$$C = \sum_i V_i \lambda_i \tag{SI-14}$$

where $i$ indicates the bronchiolar ($B$), laryngeal ($L$) or oral ($O$) mode. Explicitly,

$$C = V_{\mathrm{B}}\lambda_{\mathrm{B}} + V_{\mathrm{L}}\lambda_{\mathrm{L}} + V_{\mathrm{O}}\lambda_{\mathrm{O}}. \tag{SI-15}$$

Studies such as Coleman et al. (2021) report that a percentage ($p^*$) of the number of virus copies detected is present in particles with diameter ($d_{\mathrm{p}}$) smaller than a threshold particle diameter ($d_{\mathrm{p}}^*$; [µm]), where

$$p^* = \frac{C(d_{\mathrm{p}} \leq d_{\mathrm{p}}^*)}{C}. \tag{SI-16}$$

For example, in the case of Coleman et al. (2021), 85% of the gene copies were detected in particles smaller than 5 µm, and this can be interpreted as the ratio of total copy numbers $p^* = 0.85$ for threshold diameter of $d_{\mathrm{p}}^* = 5$ µm.

We hypothesise that the implication of this empirical evidence is that there is a higher viral load in the B and L modes ($\lambda_{\mathrm{BL}} = \lambda_{\mathrm{B}} = \lambda_{\mathrm{L}}$) compared with the viral load of the O mode ($\lambda_{\mathrm{O}}$). Here, we solve the ratio in viral loads ($r$) required to explain the findings of the experimental studies, where

$$r = \frac{\lambda_{\mathrm{BL}}}{\lambda_{\mathrm{O}}}. \tag{SI-17}$$

Combining SI-17 and SI-15 gives

$$C = \left( V_{\mathrm{B}} + V_{\mathrm{L}} + \frac{V_{\mathrm{O}}}{r} \right) \lambda_{\mathrm{BL}} \tag{SI-18}$$

and this can also be written as the virus copies in particles with diameter less than a threshold diameter as,

$$C(d_{\mathrm{p}} \leq d_{\mathrm{p}}^*) = \left( V_{\mathrm{B}}(d_{\mathrm{p}} \leq d_{\mathrm{p}}^*) + V_{\mathrm{L}}(d_{\mathrm{p}} \leq d_{\mathrm{p}}^*) + \frac{V_{\mathrm{O}}(d_{\mathrm{p}} \leq d_{\mathrm{p}}^*)}{r} \right) \lambda_{\mathrm{BL}} \tag{SI-19}$$

Then, combining SI-18, SI-19 and SI-16 and solving for $r$,

$$r = \frac{V_{\mathrm{O}}(d_{\mathrm{p}} \leq d_{\mathrm{p}}^{*}) - V_{\mathrm{O}}p^{*}}{V_{\mathrm{BL}}p^{*} - V_{\mathrm{BL}}(d_{\mathrm{p}} \leq d_{\mathrm{p}}^{*})}, \tag{SI-20}$$

where $V_{\mathrm{BL}} = V_{\mathrm{B}} + V_{\mathrm{L}}$. This equation can be used to determine the ratio of the viral load in the B and L modes to that in the O mode, given $p^{*}$ and $d_{\mathrm{p}}^{*}$ and assuming a exhaled particle size distribution.

The final step is to make a judgment on whether the particles measured by an experiment represent the equilibrium particle size distribution or the particle size distribution at the mouth. In the case of Coleman et al. (2021), we judge that the particles are at equilibrium (i.e. they have been dried) as a result of dilution of the exhaled breath with room air and transport of the aerosol through a sampling system prior to collection of particles with an aerodynamic diameter greater than 5 µm via impaction. We note that in the original paper describing this apparatus, the exhaled air is diluted with 'humid' air that is delivered as a sheath flow around a sampling cone and that the individual is located in a booth with a humidifier (McDevitt et al., 2013). However, these measures do not appear in the image shown as Figure 1 in Coleman et al. (2021).

The effect of particle drying is to reduce the total particle volume and thus 'concentrate' the viral load in the B and L modes (recall that the O mode is not corrected for drying by Johnson et al. (2011)). Thus, to correct $r$ to account for drying of particles and to calculate its value for particles as they exit the mouth, it should be multiplied by the ratio of the total particle volume in the B and L modes at equilibrium to the volume at the mouth,

$$r_{\mathrm{Mo}} = r_{\mathrm{Eq}} \frac{V_{\mathrm{BL,Eq}}}{V_{\mathrm{BL,Mo}}}, \tag{SI-21}$$

where the subscripts 'Eq' and 'Mo' represent the particle size distribution at equilibrium and at the mouth, respectively. Values for $V_{\mathrm{O}}$= $9.46\times10^{3}$ µm$^{3}$ cm$^{-3}$, $V_{\mathrm{O}}(d_{\mathrm{p}} \leq d_{\mathrm{p}}^{*})$, $V_{\mathrm{BL}}$= 0.217 µm$^{3}$ cm$^{-3}$ and $V_{\mathrm{BL}}(d_{\mathrm{p}} \leq d_{\mathrm{p}}^{*})$ corresponding to the equilibrium particle size distribution shown in Figure 3(b)(ii) (main text) for speaking are shown graphically in Figure SI-1. For particles exhaled during speaking, $\frac{V_{\mathrm{BL,Eq}}}{V_{\mathrm{BL,Mo}}} = 0.1251$. In Figure SI-2 we show $r$ as a function of $p^{*}$ for $d_{\mathrm{p}}^{*}$ equal to 5µm and 10µm.

Taking the results from Coleman et al. (2021), where $d_{\mathrm{p}}^{*} = 5$µm and $p^{*} = 0.85$, we can find that $V_{\mathrm{O}}(d_{\mathrm{p}} \leq d_{\mathrm{p}}^{*}) \approx 0$ µm$^{3}$ cm$^{-3}$ and $V_{\mathrm{BL}}(d_{\mathrm{p}} \leq d_{\mathrm{p}}^{*}) = 0.197$ µm$^{3}$ cm$^{-3}$. Thus, $r \approx 6\times10^{5}$, i.e. given assumptions on the particle size distribution, the viral load in the B and L modes must be $6\times10^{5}$ times the viral load in the O mode.

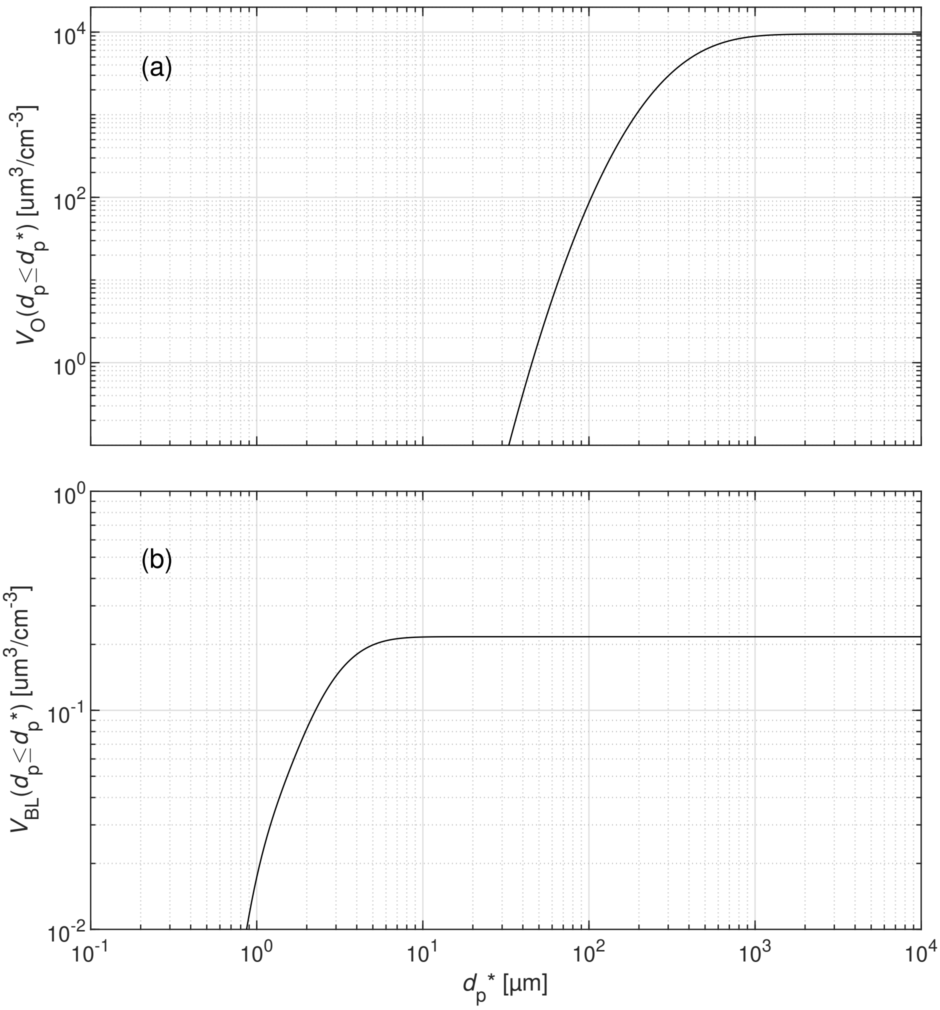


Figure SI-1: Cumulative particle volume concentration for the (a) O-mode and (b) sum of B and L modes for the equilibrium particle size distribution reported by Johnson et al. (2011).

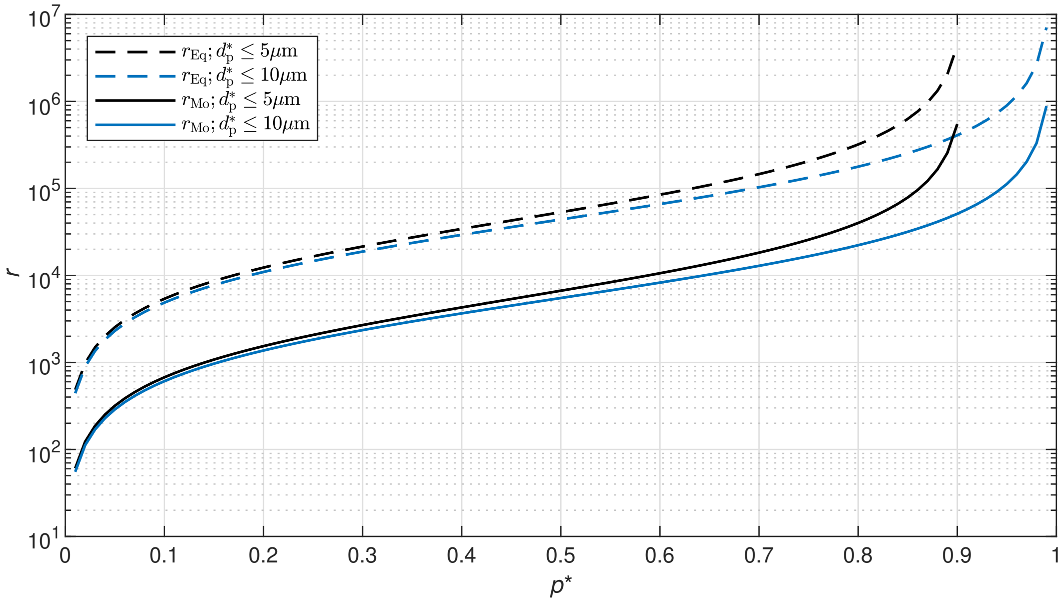


Figure SI-2: Ratio of viral loads in the B and L modes relative to that in the O mode ($r$) as a function of the the percentage ($p^*$) of virus copies detected for particles with diameter less than a threshold diameter ($d_{\mathrm{p}}^*$) equal to 5μm and 10μm.
